# Multi-Centre Randomised Controlled Feasibility Testing of a Physical Activity Micropattern Intervention Among Socioeconomically Diverse Women

**DOI:** 10.1101/2025.10.28.25339013

**Authors:** Nicholas A. Koemel, Maria Bissett, Gemma C. Ryde, Matthew Ahmadi, Kristina Atsiaris, Iris Chen, Armando Teixeira-Pinto, Sam Liu, Carol Maher, Kar Hau Chong, Adrian Bauman, Mouna Sawan, Cecilie Thøgersen-Ntoumani, Cindy M. Gray, Mark Hamer, Jason M.R. Gill, Emmanuel Stamatakis

**Affiliations:** Mackenzie Wearables Research Hub, Charles Perkins Centre, The University of Sydney, Sydney, New South Wales, Australia; School of Health Sciences, Faculty of Medicine and Health, The University of Sydney, Sydney, New South Wales, Australia; School of Cardiovascular and Metabolic Health, University of Glasgow, UK; Sydney School of Public Health, University of Sydney, Camperdown, New South Wales, Australia; Centre for Kidney Research, The Children’s Hospital at Westmead, New South Wales, Australia; School of Exercise Science, Physical & Health Education, University of Victoria, Canada; Alliance for Research in Exercise, Nutrition and Activity, Allied Health and Human Performance, University of South Australia, Australia; School of Social Sciences, Faculty of the Arts, Social Sciences and Humanities, University of Wollongong, Wollongong, New South Wales, Australia; Sydney School of Public Health, University of Sydney, Sydney, New South Wales, Australia; School of Pharmacy, Faculty of Medicine and Health, The University of Sydney, Sydney, New South Wales, Australia; Danish Centre for Motivation and Behaviour Science (DRIVEN), Department of Sports Science and Clinical Biomechanics, University of Southern Denmark, Odense, Denmark; School of Sport, Exercise, and Rehabilitation Sciences, University of Birmingham, United Kingdom; School of Social and Political Studies, University of Glasgow, UK; Institute of Sport Exercise and Health, Division of Surgery and Interventional Sciences, UCL, United Kingdom; University College London Hospitals NIHR Biomedical Research Centre, London, UK

**Keywords:** Physical activity, micropatterns, incidental physical activity, behaviour change, intervention, feasibility study

## Abstract

**Background:** Most physical activity interventions adopt a one-size-fits-all approach, often developed and tested in higher socioeconomic groups, with little consideration of barriers experienced by women from lower socioeconomic backgrounds. Micropatterns, defined as brief bursts of moderate-to-vigorous incidental physical activity lasting under three minutes, represent a form of everyday movement that requires minimal time, equipment, or planning, making them a promising target for intervention. This randomized controlled study evaluated the feasibility, acceptability and potential effectiveness of a co-designed wearable-based micropatterns intervention aimed at increasing moderate-to-vigorous physical activity (MVPA) among socioeconomically diverse women.

**Methods:** A multi-centre, single-blinded, 6-week randomised controlled feasibility study was conducted among women from Sydney, Australia and Glasgow, UK aged ≥30 years who self-reported no leisure time physical activity and could comfortably participate in activities of daily living. To ensure socioeconomic diversity, we used quota-based recruitment with area-level and individual-level indicators of socioeconomic status. Participants were randomised to one of three intervention arms; 1) Smartphone application only (APP); 2) smartphone application combined with a wearable device (Fitbit Inspire 3; W-APP); and 3) smartphone application and wearable device, supplemented by a social community component that included two in-person workshops and online forums (W-APP-C). Feasibility was assessed as completion rates of the study and follow-up surveys were conducted to assess the acceptability of the intervention. Wrist-worn accelerometry was collected at pre- and post-intervention to assess the potential effectiveness of the intervention.

**Results:** Forty-three participants (mean age (SD); 54.8 (10.7) years) were recruited across both sites (APP n = 13; W-APP n = 17; W-APP-C n = 13). When taking into consideration individual-level indicators of socioeconomic status including income, number of dependents, and educational attainment, there was diverse representation across low (20.9%), medium (48.8%), and high (30.2%) socioeconomic groups. Completion of the 6-week intervention was high across both intervention sites and all three trial arms (86%). Qualitative feedback from both trial sites indicated high satisfaction, the majority of the participants thought the intervention was enjoyable (97.9%), user-friendly (69.9%), the wearable technology increased motivation to do activity (87.5%), and the community workshops were beneficial (75%). Across all participants, there was an average daily increase of 9.8 (27.8) min/day MVPA following the intervention. The greatest increase in MVPA was observed in the W-APP-C group (15.5 (23.1) min/day), followed by the APP group (11.4 (25.5) min/day), and the W-APP group 2.6 (32.1) min/day.

**Conclusions:** This multi-centre randomised controlled feasibility trial highlights the feasibility and acceptability of a wearable-based micropattern intervention in women from socioeconomically and culturally diverse backgrounds.

## INTRODUCTION

Physical inactivity is a major risk factor for non-communicable diseases (NCD), including cardiovascular disease (CVD), type 2 diabetes, and many types of cancer^1–3^. Both clinical and public health practice have traditionally focused on promoting longer bouts of leisure-time exercise (e.g. sports, gyms, exercise classes), a domain that is characterised by wide gender inequalities in participation^4^, primarily due to significant resource-related barriers including a lack of time, equipment, or access to facilities, more frequently encountered by socioeconomically disadvantaged communities^5^. Incidental physical activity^6–9^, defined as non-exercise activities that are done as part of daily living (e.g. carrying shopping, housework, using the stairs) has important feasibility advantages, as it overcomes most traditional barriers to exercise, such as poor access to facilities^5^. This also has major public health relevance as only 20% of adults regularly participate in leisure-time exercise, with incidental movement being the predominant source of physical activity for the majority of adults^10^. This is particularly relevant for women who have 18-58% lower participation rates in leisure-time sports compared to men^11,12^.

Recent evidence has revealed that the total amount of lifestyle (incidental) physical activity (i.e. transportation, work or domestic activities done as part of daily routines outside the exercise domain) shows steep beneficial associations with CVD mortality and major adverse cardiovascular events^8,13,14^. In particular, short bouts of vigorous intermittent lifestyle physical activity (VILPA) can reduce mortality from cardiovascular disease and some forms of cancer. As little as 4-6 minutes per day of VILPA^14,15^, undertaken in bouts of up to 1-2 minutes in duration, has been shown to be especially beneficial and appears highly feasible to improve health among individuals who do little to no planned exercise. In a recent UK Biobank study including 22,000 UK adults, VILPA^15^ has also demonstrated a uniquely higher cardioprotective association in women compared to men. There have been analogous cardioprotective findings for moderate-to-vigorous intermittent lifestyle physical activity (MV-ILPA), undertaken in bouts up to three minutes in duration^8^. Such “micropatterns” of physical activity accrued through very short bursts do not require the time, equipment or preparation that typically accompanies structured forms of leisure-time physical activity or planned exercise and therefore such interventions may be more accessible to socioeconomically disadvantaged populations. For example, these may include activities such as brisk walking, taking the stairs, outdoor gardening, carrying heavy loads during household chores, or energetic playing with children^8,16^. However, incidental physical activity, including amounts accrued through moderate-to-vigorous micropatterns, may have unique barriers and facilitators that need to be explored to design interventions suitable for community integration. Limited evidence exists for the feasibility of using digital tools, including smartphone applications and wearable devices to deliver interventions aimed at improving individual-level micropattern profiles.

This study describes a 6-week feasibility study of a micropatterns intervention co-designed with socioeconomically diverse community members in the UK and Australia^17^ to assess the feasibility, acceptability, and potential effectiveness of such an intervention. We also aim to test core features of importance we identified during the co-design process, including the importance of wearable devices and social community involved in the delivery of the intervention. The findings will inform the feasibility of conducting large scale randomised trials of physical activity micropattern interventions, including the use of wearables and smartphone applications, across socioeconomically diverse women.

## METHODS

### Feasibility Trial and Study Design

This multi-centre randomised controlled feasibility trial was conducted in the UK and Australia and builds on prior co-design work. In our preparatory micropatterns co-design work^17^, three experience-based workshops (Australia: n = 31; UK: n = 19) explored perceptions of micropatterns, barriers and facilitators, and preferences for intervention delivery. Insights from this process informed the development of the 6-week intervention tested in the present study, which examined the feasibility of using smartphones and consumer-grade wearables to promote behaviourally sustainable micropatterns, supported by pre-recorded weekly educational content. The trial was conducted independently in Glasgow, UK, and Sydney, Australia to improve generalisability across cultural, environmental, and geographical contexts.

### Recruitment and Eligibility

Participants were recruited from March 2025 through September 2025 via a combination of social media advertisements through community contacts and flyers placed in community centres, libraries, and organisations providing services to those in disadvantaged communities. Eligible participants were females aged 30 years and above who were able to comfortably participate in activities of daily living. We aimed to recruit 50% of the sample from more disadvantaged communities, specifically targeting the two lowest quintiles of area-based socioeconomic status indices (i.e., Socioeconomic Indexes for Areas (SEIFA)^18^ score in Australia and Scottish Index of Multiple Deprivation (SIMD)^19^ in the UK). Individual-level indicators of socioeconomic status (SES) including household income, number of dependents and education were assessed also explored to ensure socioeconomic diversity. Socioeconomic indicators were categorised using rankings from 1 to 5 based on predefined criteria. Income was equivalised to the number of household dependents^20^. A composite socioeconomic status score was then calculated, with scores ranging from 1.0–2.5 classified as low, >2.5–3.5 as medium, and >3.5–5.0 as high.

Participants were excluded from the study if they 1) reported participating in any leisure-time exercise or recreational walks lasting for at least 10 minutes per day; 2) reported engaging in vigorous physical activity as a part of work that causes large increases in breathing or heart rate (e.g., carrying or lifting heavy loads, digging or construction work); 3) did not satisfy the Physical Activity Readiness Questionnaire (PARQ+) criteria and were advised by a physician to not participate; or 4) were unable or unwilling to use smartphone apps or wearable technology (e.g., Fitbit or analogous device). All participants provided either written or digital consent to participation in this study, and the project was approved by ethics committees in both the UK (University of Glasgow, College of Medicine, Veterinary and Life Sciences College Ethics Committee: 200240234) and Australia (The University of Sydney, Human Research and Ethics Committee: HEOO1729).

### Randomisation and Blinding

Fully consented participants were randomised (1:1:1) to a single-blinded intervention consisting of three arms including 1) a smartphone application only (APP); 2) a smartphone application plus wearable device (Fitbit Inspire 3; W-APP); or 3) a smartphone application and wearable device supplemented by social workshops and community forums (W-APP-C). Randomisation was performed for all consented participants using a computer-generated allocation to ensure concealment at each site. Participants were blinded to the intervention arm and study design and enrolled as a cohort for W-APP-C and on a rolling basis for the APP and W-APP arms.

### Baseline and follow-up measures

At baseline, all participants across all three groups joined a 30-minute one-on-one in-person information session to answer a self-report questionnaire and receive an overview of the intervention including the technology set-up. Physical activity levels were assessed at baseline (week 0) and follow-up measurement (week 7) using a wrist-worn research-grade accelerometer (UK: Actigraph wGT3X-BT; Australia: Axivity AX3) to assess changes in micropatterns following a 7-day wear time procedure^6–8,13–15,21^. To be included in our analyses, participants must have worn the device at least three valid monitoring days (>16 hours), including at least one weekend day. All accelerometry data were processed using a validated two-stage random forest classifier^6–8,13,14,21^. Additional details regarding the validation and performance of the algorithm are provided in the **Supplementary Methods 1**.

### The Micropattern Intervention

Throughout the preparatory co-design process^17^, we developed a bespoke smartphone application for delivery of intervention materials. The application was available on both android and IOS devices developed using Pathverse Platform^22^. The application enables loading of pre-recorded information sessions regarding the benefits of micropatterns, motivational tips, and goal-setting suggestions. The intervention for all groups included brief 3-10-minute educational videos providing an overview of topics of interest determined through previous conducted co-design workshops^17^ with each week mapped incrementally to address areas of capability, opportunity, and motivation^23^. Based on varying levels of physical fitness and motivation, participants were encouraged to set an initial personalised goal (i.e., daily number of bouts) and incrementally increase their goal over the course of the 6-week feasibility study. Each week, the participants received prompts to self-select an incrementally higher micropattern goal from the previous week. We defined micropattern bouts using minute-by-minute stepping cadence data obtained via the Fitbit API, which is integrated into the Pathverse platform. The Pathverse platform^22^ automatically retrieves timestamped step count data (steps per minute) and allows users to define and visualize their own micropattern VILPA bouts. In this study, we defined a micropattern bout as a period with a stepping cadence >100 steps per minute or stairs climbed that occurred in a duration of <3 minutes to satisfy that they were accrued as brief moderate-to-vigorous intermittent bursts^6,8,14,15,24^.

For the W-APP group, each participant was provided a consumer-grade wearable device (Fitbit Inspire 3) for the duration of the intervention with linkage to the intervention application for micropattern estimation. For the W-APP-C group, participants received the consumer grade wearable device in addition to two additional group information sessions in weeks 1-2 of the intervention and a community forum to enable the participants in this arm to socialise. Further detail of the intervention procedure including the development of the smart phone application and week-by-week topics is provided in the **Supplementary Methods 2**.

### Micropattern Intervention Feasibility and Acceptability

The primary outcome, which included feasibility and acceptability of the intervention, was determined by the completion rate of participants and the participant feedback at the end of the intervention. We conducted an assessment of the acceptability and usability of the intervention tools using a self-reported 26-item acceptability survey once completing the study including items such as usability, need for more information, participant perspectives on the importance of the wearable device and community elements of the intervention. Changes in MVPA following the intervention were also explored to determine the most effective mode of intervention delivery. We also evaluated the level of micropatterns (MV-ILPA and VILPA), light physical activity, sedentary behaviour, and steps per day pre- and post-intervention using the research-grade accelerometry data.

### Sample Size and Analysis

The sample size was based on recommendations that groups of between 12 and 30 are sufficient for pilot studies to determine the impact of the intervention on key outcome measures in order to estimate parameters necessary to design a fully-powered trial, and to enable estimation of recruitment/retention parameters with sufficient precision^25–27^. The group-wise differences and total intervention effect were examined as the mean difference (MD) ± standard deviation (SD). All data processing and analyses were undertaken using R (version 4.4.2). We followed the CONSORT checklist for pilot and feasibility trials (**Supplementary Table 1**).

## RESULTS

### Baseline Participant Characteristics

Of the participants randomised to the three intervention arms (mean (SD); Australia: 52.9 (11.2) years, UK: 52.9 (11.2) years), the participants reflected a diverse range of ethnicities including White (60.5%), Asian (23.3%), African (14.0%) and other backgrounds (2.2%) (**Table 1**). When taking into consideration individual-level indicators of socioeconomic status including income, number of dependents, and educational attainment, there was diverse representation across low (20.9%), medium (48.8%), and high (30.2%) socioeconomic status.

**Table 1.** Baseline participant characteristics^1^.

|  | Sydney, Australia<br>N = 21 | Glasgow, UK<br>N = 22 |
| --- | --- | --- |
| **Age** (mean ± SD) | 52.9 ± 11.2 | 56.6 ± 10.1 |
| **Ethnicity** (n (%)) |  |  |
| Asian | 8 (38.1%) | 2 (9.1%) |
| African | 0 (0%) | 6 (27.2%) |
| White | 13 (61.9%) | 13 (59.1%) |
| Other | 0 (0%) | 1 (4.5%) |
| **Education** (n (%)) |  |  |
| No formal education | 0 (0%) | 0 (0%) |
| Secondary School | 1 (4.8%) | 3 (13.6%) |
| Higher education | 3 (14.3%) | 8 (36.4%) |
| University Education or higher | 17 (81.0%) | 11 (50.0%) |
| **Household Income Range** (n (%)) |  |  |
| <\$20,000 | 1 (4.8%) | 0 (0%) |
| \$20,000-\$39,999 | 1 (4.8%) | 8 (34.8%) |
| \$40,000-\$59,999 | 5 (3.8%) | 6 (26.1%) |
| \$60,000-\$79,999 | 0 (0%) | 0 (0%) |
| \$80,000-\$99,999 | 4 (19.0%) | 5 (21.7%) |
| >\$100,000 AUD | 9 (42.9%) | 0 (0%) |
| Prefer not to say | 1 (4.8%) | 4 (17.4%) |
| **Self-reported Health Status (0-100)** | 69.29 ± 22.0 | 73.3 ± 22.0 |
| **Number of dependents** | 0.81 ± 0.98 | 0.55 ± 1.14 |
| **Deprivation Index**<br>**SEIFA (regional score) (n (%))** |  |  |
| 1-5 | 14 (14.3%) | — |
| 6-10 | 18 (90.4%) | — |
| **Deprivation Index SIMD**<br>**(area-level Score)** | — | 15 (68.2%) |
|  | — | 7 (31.8%) |
| **Individual Level Indicators of Disadvantage** |  |  |
| Low SES, n (%) | 4 (19%) | 5 (22.7%) |
| Lower-middle SES, n (%) | 13 (62%) | 8 (36.4%) |
| High SES, n (%) | 4 (19%) | 9 (40.9%) |

<sup>1</sup>All values are presented as mean ± standard deviation (SD). Individual level indicators of socioeconomic status (SES) including household income, number of dependents and education. Socioeconomic indicators were categorised using rankings from 1 to 5 based on predefined criteria. A composite socioeconomic status score was then calculated, with scores ranging from 1.0–2.5 classified as low, >2.5–3.5 as medium, and >3.5–5.0 as high. SEIFA, Australian Socioeconomic Indexes for Areas; SIMD, Scottish Index of Multiple Deprivation.

### Feasibility and Acceptability of the Micropattern Intervention

A total of 22 participants in Sydney (100%) and 18 participants in Glasgow (81.2%) completed the intervention (**Figure 1**). Study withdrawal rates were 23.5% in the APP arm (n = 4) and 15.4% in the W-APP arm (n = 2), while no participants withdrew from the W-APP-C arm. All participants from both sites successfully completed the pre- and post-intervention accelerometry assessment. Across all three groups in both countries, there was a high level of participant satisfaction with the intervention (97.9%; **Table 2**). Participants in the W-APP-C arm had a higher overall agreement for using the intervention application (56.3% vs. 35.6% across other intervention arms) and a greater perception that the application had a positive impact on physical activity levels (50.0% vs. 28.9% across other intervention arms). Participants across all groups and sites strongly agreed that the Fitbit motivated them to be more active than using the application on its own (87.5% across other intervention arms). All participants in the W- APP-C reported satisfaction from completing the social workshops (100%) and its role in helping them understand how to incorporate micropatterns in everyday living (75%).

**Figure 1.**
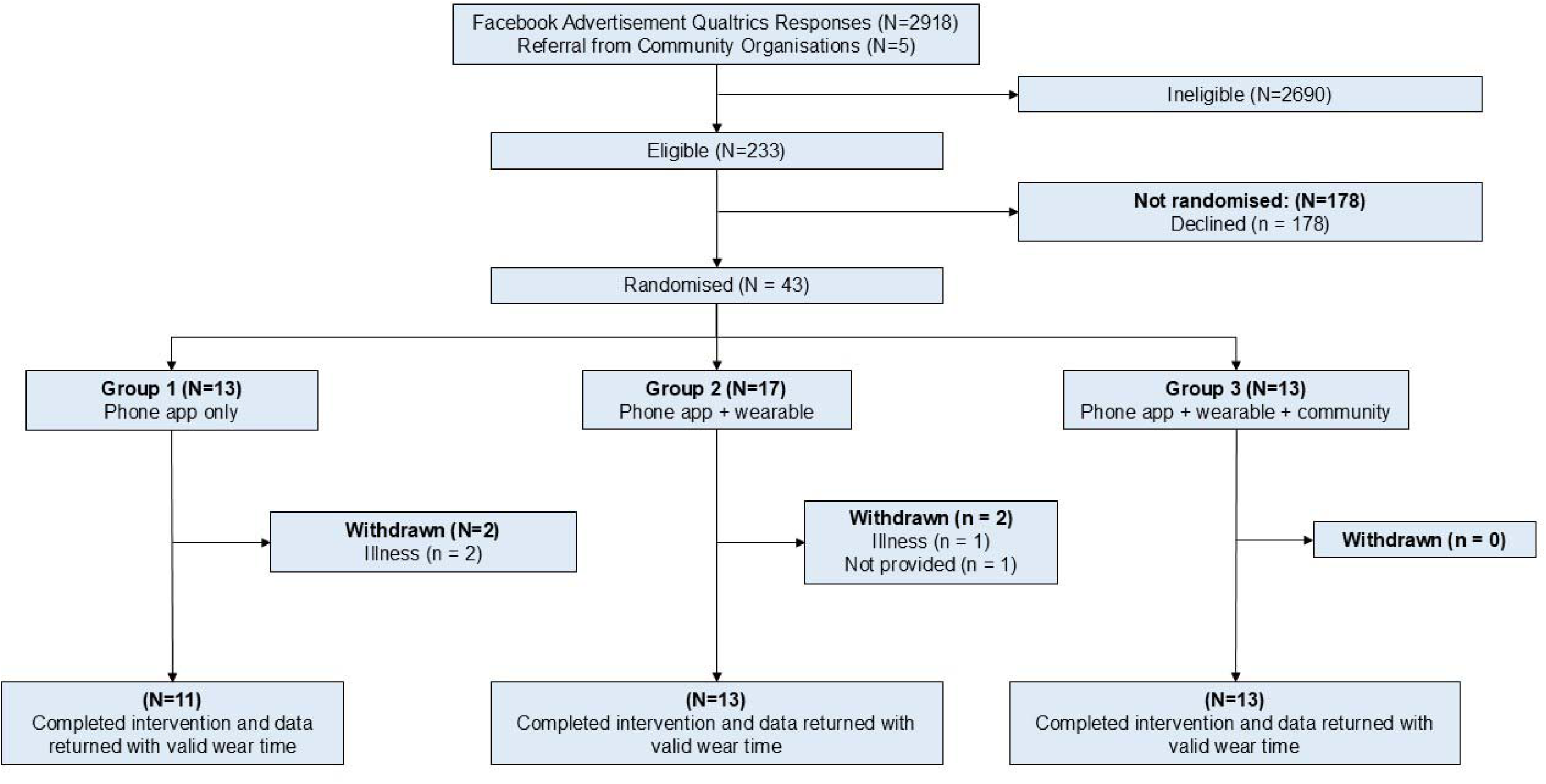
Study Participant Flow Chart.

**Table 2.**
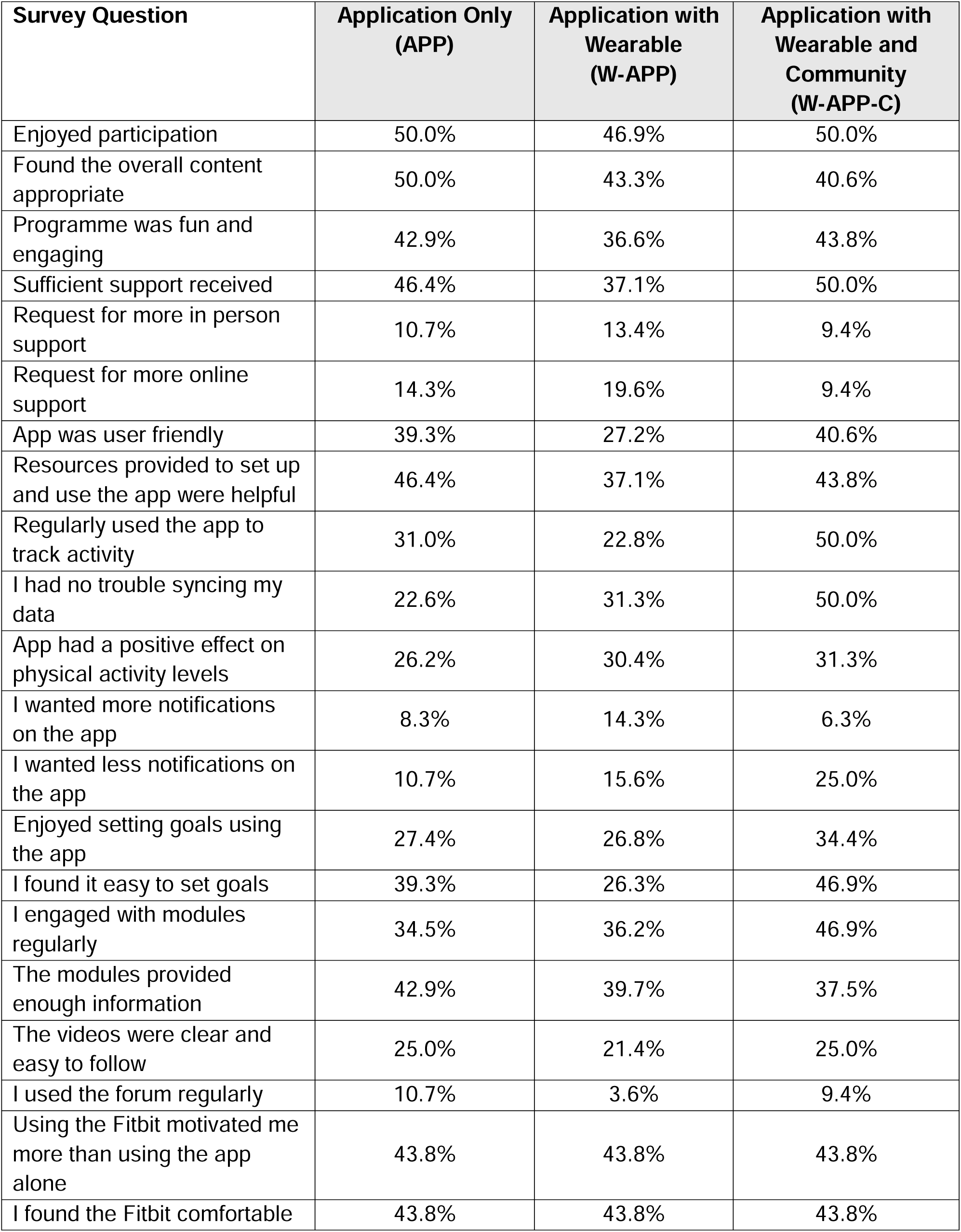
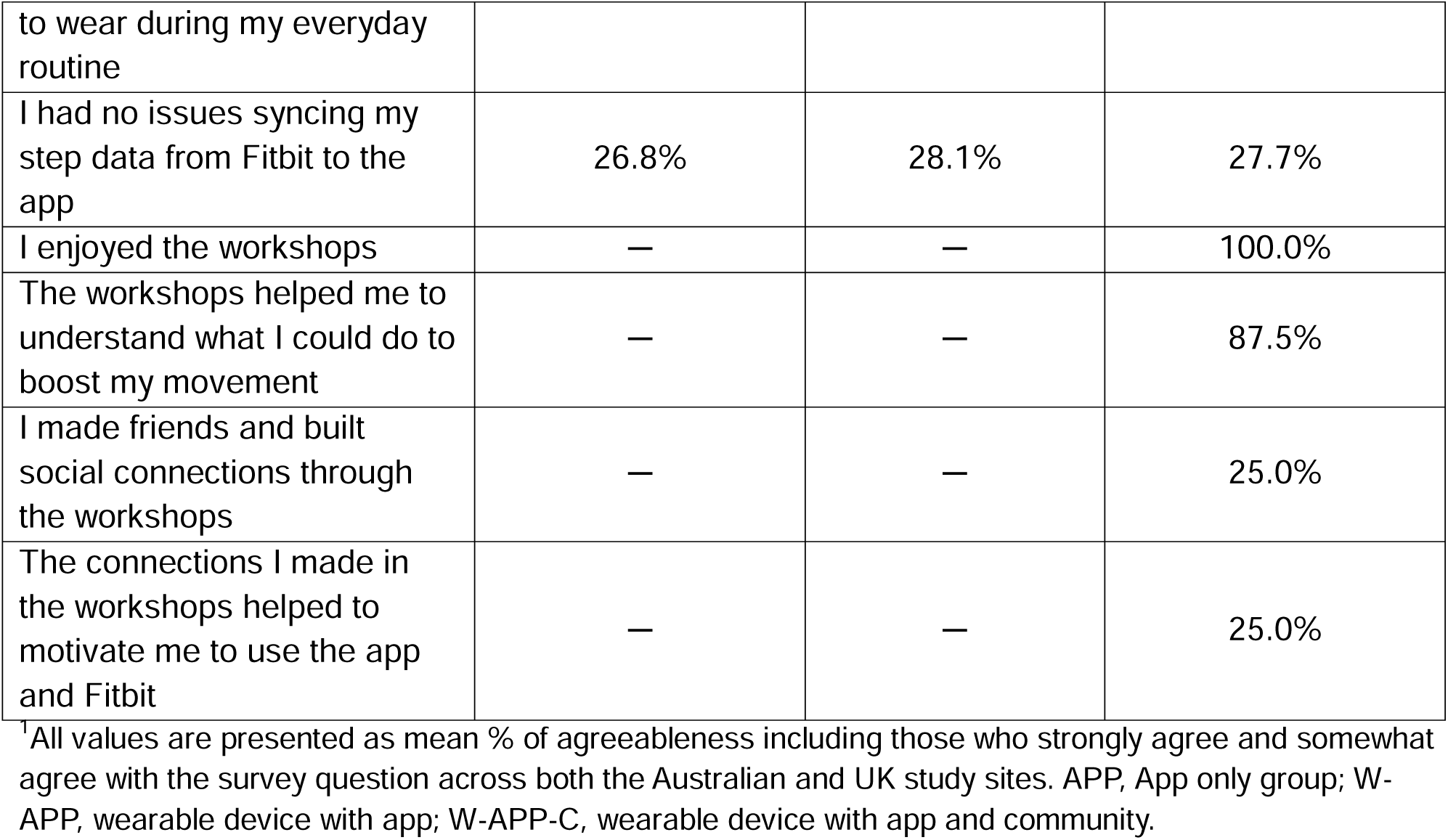
Micropattern interventions survey of acceptability and usability^1^.

### Physical activity and Sedentary Behaviour

Across all three groups, there was a post-intervention improvement in MVPA of 9.8 (27.8) minutes per day. The mean daily MVPA change was greatest in W-APP-C which had an increase of 15.5 (23.1) minutes per day (**Table 3**). The APP group (i.e., no wearable or community), had the second greatest improvements of 11.4 (25.50) minutes per day increase. The W-APP who received a wearable device but no social or community element had a more modest improvement in physical activity following the intervention including 2.6 (32.1) minutes of additional MVPA per day.

**Table 3.** Micropattern interventions differences following the intervention^1^.

|  | **Application Only**<br>**(APP)** |  |  | **Application with Wearable**<br>**(W-APP)** |  |  | **Application with Wearable and**<br>**Community**<br>**(W-APP-C)** |  |  |
| --- | --- | --- | --- | --- | --- | --- | --- | --- | --- |
| **Behaviour** | Pre | Post | MD | Pre | Post | MD | Pre | Post | MD |
| Total MVPA, min/day | 26.5 (27.4) | 37.9 (23.4) | 11.4 (25.50) | 40.9 (22.8) | 43.5 (40.0) | 2.6 (32.1) | 28.5 (14.2) | 44.0 (30.9) | 15.5 (23.1) |
| VPA, min/day | 2.2 (2.4) | 3.5 (4.3) | 1.3 (3.7) | 4.0 (3.7) | 3.5 (5.3) | -0.5 (4.44) | 3.1 (3.1) | 3.5 (4.33) | 0.4 (3.73) |
| MPA, min/day | 24.3 (25.4) | 34.4 (32.8) | 10.1 (29.8) | 36.9 (21.3) | 40.0 (34.9) | 3.1 (28.6) | 25.4 (12.9) | 40.5 (31.9) | 15.1 (24.1) |
| VILPA, min/day | 2.2 (2.4) | 3.5 (4.3) | 1.3 (3.73) | 3.6 (2.7) | 3.5 (5.3) | -0.1 (4.44) | 3.1 (3.1) | 3.1 (3.7) | 0.0 (3.43) |
| MV-ILPA, min/day | 24.5 (25.7) | 32.3 (29.7) | 7.8 (27.92) | 36.3 (20.9) | 38.5 (36.8) | 2.2 (29.4) | 26.1 (13.7) | 38.1 (28.1) | 12.0 (21.9) |
| Light physical activity, min/day | 305.0 (73.1) | 321.3 (57.3) | 16.3 (66.62) | 297.0 (62.5) | 286.0 (83.1) | -11.0 (73.0) | 386.0 (56.1) | 415.0 (102.0) | 29.0 (79.6) |
| Sedentary time, min/day | 610.5 (106.5) | 522.0 (97.3) | 88.5 (102.21) | 569.0 (85.3) | 546.9 (124.0) | -22.1 (104.2) | 546.1 (90.9) | 454.5 (119.7) | -91.6 (105.4) |
| Total steps per day | 9,448 (3,398) | 10,123 (3,4747) | 675 (3437) | 11,057 (3,505) | 11,260 (4,589) | 203 (4,047) | 11,434 (3,227) | 13,156 (4,886) | 1,722 (4,080) |

<sup>1</sup>All values are presented as mean ± standard deviation (SD) derived from research-grade accelerometers. Abbreviations: APP, App only group; W-APP, wearable device with app; W-APP-C, wearable device with app and community; MVPA, moderate-to-vigorous physical activity; VPA, vigorous physical activity; MPA, moderate physical activity; MD, mean difference, VILPA, vigorous intermittent lifestyle physical activity; MV-ILPA, moderate-to-vigorous intermittent lifestyle physical activity; LPA, light physical activity.

Total micropatterns of physical activity followed a similar grouping for MV-ILPA including an additional 7.8 (27.92), 2.2 (29.4), and 12.0 (21.9) minutes per day for APP, W-APP, and W-APP-C, respectively. There were modest to no changes in VILPA across the groups including 1.3 (3.73) minutes per day, -0.1 (4.44), 0.0 (3.43) for APP, W-APP, and W-APP-C, respectively. Across all three groups, there was an improvement of 11.5 (73.7) minutes per day in light physical activity and 866.7 (3,930) steps per day. Sedentary time was reduced by 67.4 (103.9) minutes per day following the intervention.

## DISCUSSION

This multi-centre randomised feasibility study of Australian and UK adults provides the first evidence supporting the feasibility of wearable-based micropatterns intervention for promoting increases in MVPA across socioeconomically diverse women. The aim of this study was to evaluate the feasibility in order to support progression to a fully powered clinical trial in this population. Our findings demonstrate promising feedback from the intervention indicated strong acceptability and usability of the intervention application and highlighted the use of the consumer-grade wearable for motivation. The high completion (86%), agreeableness, and usability of the intervention, and observed physical activity improvements following the intervention support the feasibility of testing larger, community wide interventions aimed at improving micropatterns of physical activity in women. In particular, we observed the highest completion rates and agreeableness in those who participated in intervention arm with community groups and discussion forums (W-APP-C). We also noted promising behavioural profile improvements of physical activity following the 6-week intervention, including an average increase of approximately 10 minutes greater incidental MVPA minutes per day, corresponding to almost 47% of the weekly amounts recommended by WHO^28^.

Previous physical activity interventions^29,30^ have predominantly been conducted in affluent communities, with few trials incorporating community perspectives of women or considering the barriers they face to engaging in physical activity^31^. Trials in women and disadvantaged communities^32,33^ have demonstrated disproportionately lower adherence and limited to no improvements compared to more affluent communities which may have greater accessibility to education, equipment and facilities. The emergence of digital tools^34–36^ offers a promising strategy to democratise access to physical activity education, support habit formation, and improve adherence to interventions in underserved communities.

Building on a growing body of population-level micropattern research^6–9,13,14,37^, this multi- centre feasibility intervention offers evidence for the translation of micropatterns into an intervention tailored to women and disadvantaged community members. This intervention was shaped by an extensive international co-design process^17^ to inform community perspectives on the education, feasibility, and practicality of the intervention explored in this study. Unlike previous behavioural physical activity interventions which often experience high-attrition rates of 17-28%^38,39^, our trial demonstrated a low drop- out rate of 9.1% across Australia and the UK. We also observed meaningful improvements in physical activity of approximately 10 minutes per day of incidental MVPA after only 6-weeks of an intervention.

This multi-centre randomised feasibility of micropatterns study supports the feasibility of a digital and wearable based promotion of incidental physical activity among women who do not participate in regular leisure time physical activity. By identifying opportunities to complete brief bursts of incidental moderate-to-vigorous physical activity, this intervention provides a novel approach to overcome the common barriers to structured exercise such as a lack of time, equipment, motivation,^5^ and capitalises on the integration of physical activity in everyday living. This has major public health relevance for the development of larger, community-wide interventions aimed at improving physical activity as women have 18-58% lower participation rates in leisure-time sports compared to men^11,12^. Additional large-scale randomised controlled trials of this intervention are needed to test efficacy of this approach and assess long-term behavioural adherence.

### Strengths and limitations

A major strength of this study includes the extensive multi-centre experience-based co-design process^40,41^ undertaken to design the intervention^17^. The majority of physical activity interventions globally are designed solely by researchers, and are commonly based on previously established interventions developed for other populations and localities, with only 30% employing systematic cultural or sex-specific adaptation^42^. By co-creating this intervention with women from diverse socioeconomic backgrounds, we have ensured the intervention remains practical and contextually relevant for everyday purposes. Another key strength includes the multi-centre nature of the design, which included coordinated assessment of both Australian and UK adults in Sydney and Glasgow. Considering diverse city-level and cultural characteristics has added an additional layer of cultural and built environmental generalisability to the intervention.

This study has several limitations, including the short length of the intervention and follow-up period. Due to practical considerations, we were unable to explore long-term adherence of the intervention and the role of follow-up. Throughout the intervention, some participants raised some issues with the intervention application, noting the manual need to synchronise daily activity levels. To mitigate this issue, we implemented an autonomous background system to synchronise wearable and mobile phone data. The intervention application detected micropatterns based on stepping intensity (steps per minute)^24,43^ and stairs climbed which may fail to capture other micropatterns such as incline walking or carrying a heavy load. Finally, while undertaking the recruitment for this intervention, we noted difficulties in recruiting community members from disadvantaged backgrounds. Future testing involving disadvantaged women will require additional community partners to develop trust with community members.

## CONCLUSION

In this randomised multi-centre feasibility trial, we demonstrate the feasibility and acceptability of a physical activity micropattern intervention in women from the UK and Australia. Participants in the intervention across both sites in Australia and the UK demonstrated promising increases in total incidental MVPA following the 6-week intervention. Further testing using these tools is needed to explore longer interventions to ensure the efficacy of this intervention for long-term habit formation among culturally diverse groups of women.

## Data Availability

All data produced in the present study are available upon reasonable request to the authors

## Data Availability Statement

All data produced in the present study are available upon reasonable request to the authors.

## Acknowledgements

We thank the extensive involvement of community members from Australia and the UK involved in the feasibility testing and co-design of this intervention.

