## Supplemental Document for "Multi-Centre Randomised Controlled Feasibility Testing of a Physical Activity Micropattern Intervention Among Socioeconomically Diverse Women"

**ONLINE SUPPLEMENTARY MATERIAL**

| **Page** | **Item** |
| --- | --- |
| 2 | **Supplementary Methods 1.** Micropatterns Intervention Procedure |
| 4 | **Supplementary Methods 2**. Wearable behaviour classification methods |
| 8 | **Supplemental Table 1:** CONSORT Feasibility Study Checklist |

**Supplementary Methods 1**. Wearable behaviour classification methods

**Two-stage random forest physical activity intensity and posture classification**

Incidental physical activity was classified using a validated two-stage random forest activity classifier that first classifies each 10 second window (epoch) as sedentary (lying or sitting still), stationary plus (active sitting, standing still, active standing), walking, or running (**Diagram A**)^1-3^. These activities were then classified into one of four activities including: sedentary, light, moderate, and vigorous. Walking activities (gardening, active commuting, etc) were classified by normalized gravitational units (g) where <100 milli g were classified as light intensity (<3 METs), ≥100 milli g and <400 milli g were considered moderate intensity physical activity (≥3 to <6 METs), and ≥400 milli g were considered vigorous-intensity PA (≥6 METs)^3^. All windows classified as running/high energetic activity were classified as vigorous-intensity physical activity (≥ 6 METs)^3-5^. A major advantage of this classification approach is the lower risk of possible misclassification of sporadic high-accelerations that may occur during certain stationary light activities (e.g., dishwashing)^6-8^. **Physical Activity Classification Scheme (Diagram A)**

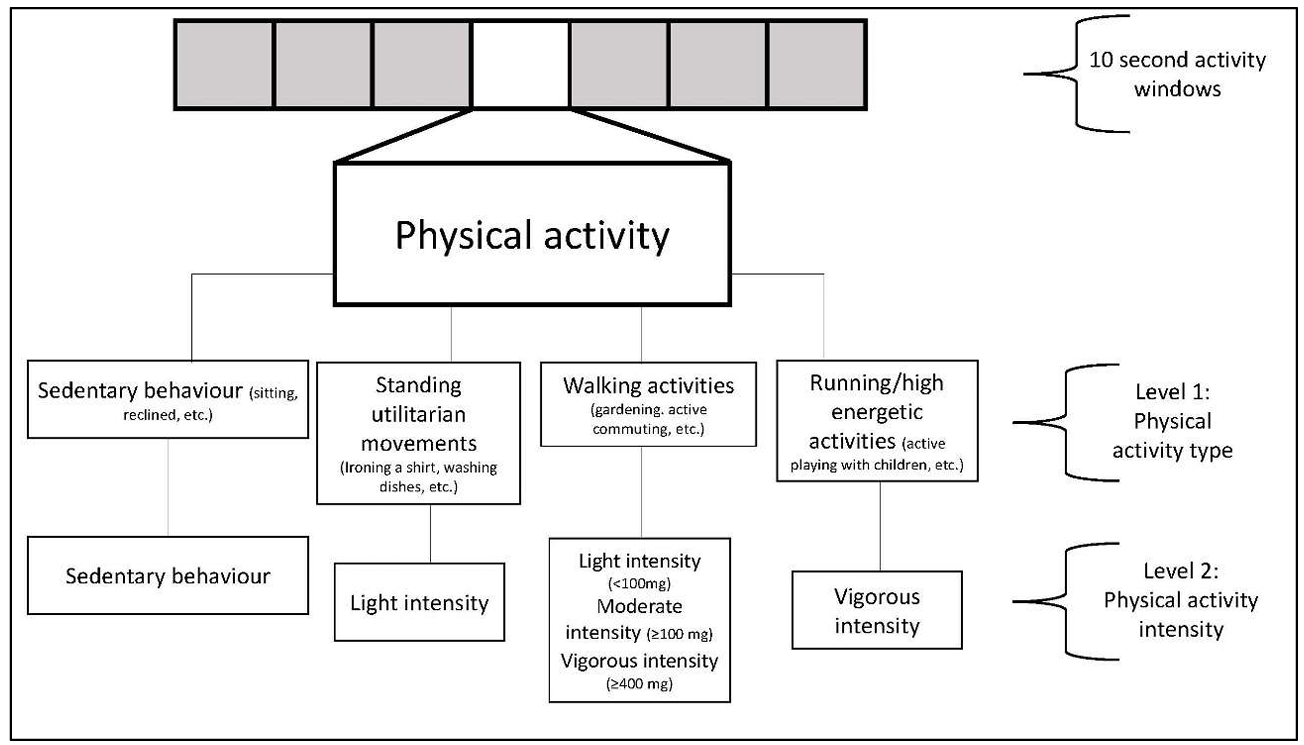

**Physical Activity Classification Performance**

The performance of this physical activity classification scheme was tested in an independent sample of 102 adults from the US^9^ and Australia^10^. This data includes direct observation measurement of 105,767 activity samples from structured and free-living activities (17,627 minutes) which were used to test the robustness and generalisability of the two-stage activity and intensity classifier. The data was collected from participant-worn or researcher-held Go-Pro video recordings. All data was imported into Noldus Observer XT software for continuous video coding. The direct observation coding generated continuous physical activity codes corresponding to the start and finish of each movement. These coded movements were then compared against the accelerometer data using the available time-stamp information. The below table includes the performance metrics across activities. Interobserver reliability was assessed by dual coding. The intraclass correlation coefficient for coding activities was 0.912 (0.866-0.942). The performance in metrics and confusion matrix for activity classification is shown below.

**Classifier Performance Metrics for Intensity in US and Australian Adults**

|  | Sensitivity | Specificity | Precision | F-score | Overall Accuracy | Weighted Kappa | Overall F-score |
| --- | --- | --- | --- | --- | --- | --- | --- |
| Sedentary | 86.5 | 93.7 | 90.5 | 88.5 |  |  |  |
| Light | 71.2 | 89.4 | 55.8 | 62.6 |  |  |  |
| Moderate | 85.4 | 96.6 | 92.7 | 88.9 |  |  |  |
| Vigorous | 95.4 | 99.4 | 94.6 | 95.0 |  |  |  |
|  |  |  |  |  | **84.6** | 0.78 | 83.8 |

Rows= ground truth; columns=predictions; bold=correct classification; all activities were free-living or simulated free-living activities.

**Confusion Matrix for Activity Classification in US and Australian Adults**

|  | Sedentary | Light | Moderate | Vigorous |
| --- | --- | --- | --- | --- |
| Sedentary | **36,904** | 5,232 | 508 | 2 |
| Light | 3,120 | **11,712** | 1,612 | 17 |
| Moderate | 502 | 4,016 | **29,528** | 526 |
| Vigorous | 226 | 17 | 214 | **9,470** |

Rows= ground truth; columns=predictions; bold=correct classification; all activities were free-living or simulated free-living activities.

**Supplementary Methods 2.** Intervention Procedures

**Rationale:** The majority of physical activity trials have been conducted in affluent high-income populations^11,12^ requiring costly intensive behavioural counselling and rarely accommodating the needs of women from diverse socioeconomic backgrounds^13^.

**Aim(s):** 1) test the feasibility of a wearables-based physical activity micropatterns intervention aimed at increasing moderate to vigorous physical activity among socioeconomically diverse UK and Australian women

**Trial Design and Setting**

This trial included a three-arm, single-blinded randomised controlled trial providing a 6-week physical activity micropattern intervention. Participants were randomised to one of three arms: 1) Smart-phone application delivery of the intervention (APP); 2) a wearable-device supported by Smart-phone app intervention alongside (Fitbit Inspire 3; W-APP ); and 3) Smart-phone application, wearable device, and social community including two additional community training workshops and community forums (W-APP-C). Participants were recruited through community organisations, social media platforms, and flyers.

**Ethics and Governance**

This trial followed the standard ethics approval process at the University of Sydney (Human Research and Ethics Committee: HEOO1729) and the University of Glasgow (College of Medicine, Veterinary and Life Sciences College Ethics Committee: 200240234) following the CONSORT guidelines for clinical trials.

**Study Population**: Women aged 30+

**Eligibility Criteria**

Participants were invited to complete a brief online Qualtrics survey which included questions regarding the following eligibility criteria:

*Inclusion Criteria*

- Adults aged 30+ years
- Able to participate comfortably in activities of daily living

*Exclusion Criteria*

- Engage in vigorous occupational activity (e.g., heavy lifting, construction).
- Do not meet the Physical Activity Readiness Questionnaire (PARQ+) survey criteria
- Unable or unwilling to use smartphone apps or wearable technology

**Recruitment and Consent**

Interested participants received a dedicated phone call to discuss the study to confirm eligibility. If an individual was eligible and interested in participating, an electronic informed consent form was provided. All participants received a reimbursement of $120 for participation in the study including compensation for transport up to $30 for each participant. Participants were recruited from the local Glasgow and Sydney areas (New South Wales) across diverse socioeconomic status (i.e., Socioeconomic Indexes for Areas (SEIFA)^14^ score in Australia and Scottish Index of Multiple Deprivation (SIMD)^15^ in the UK) prioritising recruitment from disadvantaged areas (50% of participants in the bottom two quintiles). We used recruitment quotas for each socioeconomic status quintile with target quotes across SEIFA quintiles: Quintile 1-2 (most deprived): 50%; Quintiles 3-4: 40%; Quintile 5: 10% (most advantaged).

**Randomisation and Blinding**

Fully consented participants were randomised to one of the three intervention arms following a 1:1:1 ratio. Randomisation was performed using a computer-generated allocation. The study participants were blinded to the intervention arm and the study design.

**Baseline and Follow Assessments**

Physical activity levels were assessed at baseline (week 0) and follow-up measurement (week 7) was taken using a wrist-worn research grade accelerometer (UK: Actigraph wGT3X-BT; Australia: Axivity AX3) to assess improvement in micropatterns following a 7-day wear time procedure. To be included in our analyses, participants must have worn the device at least three valid monitoring days (>16 hours), including at least one weekend day. All accelerometery data was processed using a validated two-stage random forest classifier^1,3-5,16-19^. Following the intervention (Week 7), participants completed a 26-item acceptability and usability survey.

**Interventions**

For all three arms, we developed a 10 minute introductory video and brief 3–5 minute weekly videos delivered to the participants at the beginning of each intervention week. The education was developed and themes created based on previously explored topics of interest from co-design works in socioeconomically diverse adults in the UK and Australia (publication in preparation^20^). These weekly educational modules were mapped following the established Capability, Opportunity, Motivation, Behaviour (COM-B) framework of behaviour change^21^. The development of the three arms in this study were informed by the unique input of community members during our previous micropatterns co-design workshop series^20^ supporting the adoption of wearable devices and community socialisaton during the intervention.

*Smart-phone application Development and testing*

The digital intervention was hosted on the Pathverse platform^22,23^. The Pathverse application is an iOS/Android compatible ecosystem which offers dynamic content creation and gamification features with linkage to wearable devices and internal smart-phone applications (e.g., Apple Health) while remaining compliant with data sharing laws and regulations. The Pathverse application is modular allowing flexibility for embedding pre-recorded educational videos personalised prompts and nudging. The Pathverse ecosystem also has a dedicated dashboard for participants to monitor their physical activity goals, monitor goal progression. Prior to this intervention, the Pathverse tools and resources were beta-tested at each site to ensure the functionality of the tools was adequate for the intervention. The micropatterns estimation in the application used a combination of steps per minute where a bout was defined as walking at a moderate-to-vigorous pace (>100 steps per minute^24,25^) or stair climbing for a period lasting <3 minutes as derived from the mobile phone or the Fitbit device depending on the treatment allocation arm.

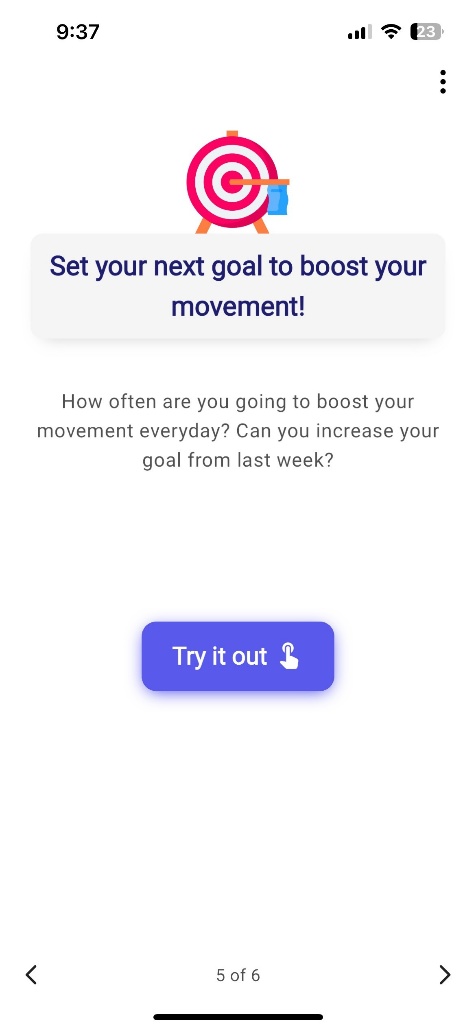

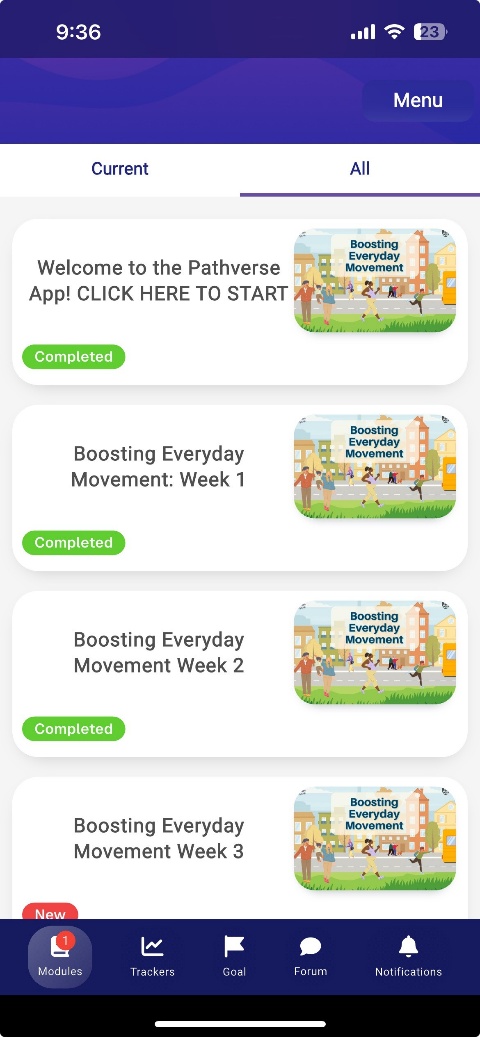

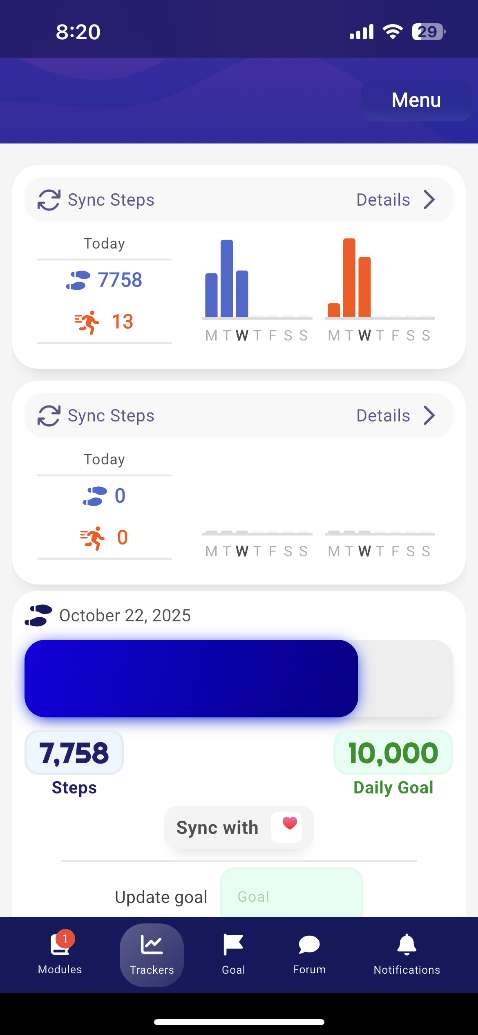

**Supplementary Methods Figure 1**. The left image shows the weekly module menu. The centre screenshot displays the activity tracking interface, indicating total steps (blue) and the number of micropattern bouts (orange). The right image shows the weekly goal-setting page, accompanied by video guidance to support incremental goal progression.

**Application Only (APP): Smart-phone application delivery of the intervention**

All participants in this arm used the smart phone micropatterns application^20^. At the launch of the intervention, participants were instructed to open the application and follow the introductory video detailing the course of the program and how to use the application. Participants then received weekly short educational videos 3-5 minutes in duration providing background information on the benefits of physical activity and instructions on setting a user-specified weekly goal. The videos and text provided each week encouraged participants to set a new goal for participants for incremental improvement each week.

**Application with Wearable (W-APP): Smart-phone delivery of the intervention alongside a wearable-device (Fitbit Inspire 3)**

All participants in this arm will use the smart phone micropatterns application including the education videos as described above for APP^20^, but with linkage to a provided Fitbit (Inspire 3).

**Application with Wearable and Community (W-APP-C): Smart-phone application, wearable device, and social community including two additional community training workshops and community forums.**

Participants in this arm had two additional group workshops in the first and second week targeting to discuss the micropatterns in more detail, including common barriers and enablers we established during our previous micropatterns co-design work^20^. To promote the social element of this intervention arm, we provided these participants with a community forum embedded in the smart-phone application to enable discussion about their experiences and reflect on new strategies to increase their physical activity. All participants in this arm will use the smart phone micropatterns application previously tested in our pilot study^20^ with linkage to a provided Fitbit (Inspire 3).

**Outcomes and data collection**

The primary outcome of the study included improvements to overall physical activity following the intervention. Primary metrics of physical activity will include total MVPA and micropatterns (VILPA/MV-ILPA). Secondary outcomes of the study included changes in light physical activity, sedentary behaviour, and total steps per day.

**Safety and Adverse Events**

Participant safety was monitored for adverse events following Good Clinical Practice procedure. No adverse events occurred during the course of this 6-week feasibility study.

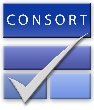
 **CONSORT 2010 checklist of information to include when reporting a pilot or feasibility trial***

| **Section/Topic** | **Item No** | **Checklist item** | **Reported on page No** |
| --- | --- | --- | --- |
| **Title and abstract** | | | |
|  | 1a | Identification as a pilot or feasibility randomised trial in the title | 1 |
|  | 1b | Structured summary of pilot trial design, methods, results, and conclusions (for specific guidance see CONSORT abstract extension for pilot trials) | 1-2 |
| **Introduction** | | | |
| Background and objectives | 2a | Scientific background and explanation of rationale for future definitive trial, and reasons for randomised pilot trial | 3 |
|  | 2b | Specific objectives or research questions for pilot trial | 3 |
| **Methods** | | | |
| Trial design | 3a | Description of pilot trial design (such as parallel, factorial) including allocation ratio | 5 |
|  | 3b | Important changes to methods after pilot trial commencement (such as eligibility criteria), with reasons | 5 |
| Participants | 4a | Eligibility criteria for participants | 5-6 |
|  | 4b | Settings and locations where the data were collected | 5 |
|  | 4c | How participants were identified and consented | 6 |
| Interventions | 5 | The interventions for each group with sufficient details to allow replication, including how and when they were actually administered | Supplementary Methods 1. |
| Outcomes | 6a | Completely defined prespecified assessments or measurements to address each pilot trial objective specified in 2b, including how and when they were assessed | Supplementary Methods 1. |
|  | 6b | Any changes to pilot trial assessments or measurements after the pilot trial commenced, with reasons | Supplementary Methods 1. |
|  | 6c | If applicable, prespecified criteria used to judge whether, or how, to proceed with future definitive trial | Supplementary Methods 1. |
| Sample size | 7a | Rationale for numbers in the pilot trial | 8 |
|  | 7b | When applicable, explanation of any interim analyses and stopping guidelines | 8 |
| Randomisation: |  |  |  |
| Sequence  generation | 8a | Method used to generate the random allocation sequence | 7 |
|  | 8b | Type of randomisation(s); details of any restriction (such as blocking and block size) | 7 |
| Allocation  concealment  mechanism | 9 | Mechanism used to implement the random allocation sequence (such as sequentially numbered containers), describing any steps taken to conceal the sequence until interventions were assigned | 7 |
| Implementation | 10 | Who generated the random allocation sequence, who enrolled participants, and who assigned participants to interventions | 7 |
| Blinding | 11a | If done, who was blinded after assignment to interventions (for example, participants, care providers, those assessing outcomes) and how | 7 |
|  | 11b | If relevant, description of the similarity of interventions | 7 |
| Statistical methods | 12 | Methods used to address each pilot trial objective whether qualitative or quantitative | 8 |
| **Results** | | | |
| Participant flow (a diagram is strongly recommended) | 13a | For each group, the numbers of participants who were approached and/or assessed for eligibility, randomly assigned, received intended treatment, and were assessed for each objective | Figure 1 |
|  | 13b | For each group, losses and exclusions after randomisation, together with reasons | Figure 1 |
| Recruitment | 14a | Dates defining the periods of recruitment and follow-up | 5 |
|  | 14b | Why the pilot trial ended or was stopped | NA |
| Baseline data | 15 | A table showing baseline demographic and clinical characteristics for each group | Table 1 |
| Numbers analysed | 16 | For each objective, number of participants (denominator) included in each analysis. If relevant, these numbers  should be by randomised group | Figure 1 |
| Outcomes and estimation | 17 | For each objective, results including expressions of uncertainty (such as 95% confidence interval) for any  estimates. If relevant, these results should be by randomised group | Table 2 |
| Ancillary analyses | 18 | Results of any other analyses performed that could be used to inform the future definitive trial | Table 2 |
| Harms | 19 | All important harms or unintended effects in each group (for specific guidance see CONSORT for harms) | NA |
|  | 19a | If relevant, other important unintended consequences |  |
| **Discussion** | | | |
| Limitations | 20 | Pilot trial limitations, addressing sources of potential bias and remaining uncertainty about feasibility | 12 |
| Generalisability | 21 | Generalisability (applicability) of pilot trial methods and findings to future definitive trial and other studies | 13 |
| Interpretation | 22 | Interpretation consistent with pilot trial objectives and findings, balancing potential benefits and harms, and  considering other relevant evidence | 12 |
|  | 22a | Implications for progression from pilot to future definitive trial, including any proposed amendments | 12-13 |
| **Other information** | | |  |
| Registration | 23 | Registration number for pilot trial and name of trial registry | NA |
| Protocol | 24 | Where the pilot trial protocol can be accessed, if available | Supplementary Methods 1 |
| Funding | 25 | Sources of funding and other support (such as supply of drugs), role of funders | 14 |
|  | 26 | Ethical approval or approval by research review committee, confirmed with reference number | 6 |

Citation: Eldridge SM, Chan CL, Campbell MJ, Bond CM, Hopewell S, Thabane L, et al. CONSORT 2010 statement: extension to randomised pilot and feasibility trials. BMJ. 2016;355. This is an Open Access article distributed in accordance with the terms of the Creative Commons Attribution (CC BY 3.0) license (<http://creativecommons.org/licenses/by/3.0/>), which permits others to distribute, remix, adapt and build upon this work, for commercial use, provided the original work is properly cited.

*We strongly recommend reading this statement in conjunction with the CONSORT 2010, extension to randomised pilot and feasibility trials, Explanation and Elaboration for important clarifications on all the items. If relevant, we also recommend reading CONSORT extensions for cluster randomised trials, non-inferiority and equivalence trials, non-pharmacological treatments, herbal interventions, and pragmatic trials. Additional extensions are forthcoming: for those and for up-to-date references relevant to this checklist, see [www.consort-statement.org](http://www.consort-statement.org).

14.	Statistics ABo. Socio-Economic Indexes for Areas (SEIFA), Australia. ABS. <https://www.abs.gov.au/statistics/people/people-and-communities/socio-economic-indexes-areas-seifa-australia/latest-release>.
